# Food Insecurity and Cardiovascular Disease Risk in Aging Women: Evidence from the Cebu Longitudinal Health and Nutrition Survey

**DOI:** 10.1101/2025.08.01.25332625

**Authors:** Rawan Ajeen, Romaniya Voloshchuk, Judith B. Borja, Linda S. Adair

**Affiliations:** Department of Nutrition, University of North Carolina at Chapel Hill, Chapel Hill, North Carolina, 27599, USA; Carolina Population Center, University of North Carolina-Chapel Hill, Chapel Hill, NC, United States; Office of Population Studies Foundation, Inc., University of San Carlos, Cebu City, Philippines

**Author notes:** Corresponding Author: Rawan Ajeen, Gillings School of Global Public Health, University of North Carolina at Chapel Hill, Chapel Hill, NC, 27599, United States. Electronic address.

**Keywords:** Philippines, Food Security, Cardiovascular Disease, Women, Low-Middle Income Country, Diabetes, Hypertension

## Abstract

This longitudinal study examined the association of food insecurity with anthropometric changes, incident hypertension and diabetes over three years in Filipino women. Data were drawn from 1,562 participants in the 2012 and 2015 Cebu Longitudinal Health and Nutrition Survey (CLHNS). Food security was measured using the 7-item Radimer-Cornell scale. Linear and logistic regression models assessed the association between 2012 food insecurity and 2015 outcomes, including BMI, waist circumference (WC), % body fat (%BF), and incident hypertension and diabetes. Higher food insecurity scores in 2012 were significantly associated with lower BMI (β = -0.14, p < 0.001), WC (β = - 0.36, p < 0.001), and %BF (β = -0.34, p < 0.001) in 2015, after adjusting for age, wealth, urbanicity, and other covariates. No statistically significant associations were found between food insecurity and incidence of hypertension (OR = 1.05, 95% CI: 0.89–1.24) or diabetes (OR = 0.92, 95% CI: 0.72–1.18). Unlike trends in high income countries (HIC), where food insecurity is linked to obesity and chronic diseases, these findings underscore the need to examine food insecurity’s effects in LMICs undergoing a nutrition transition. Future studies should include longer follow-up periods and detailed dietary assessments to clarify the role of diet as a mediator between food insecurity and disease outcomes.

## INTRODUCTION

Food security, a cornerstone of global health and sustainable development, is an integral part of human well-being and social progress. Its absence, food insecurity, is defined as the lack of regular and consistent access to sufficient, safe, and nutritious food necessary for normal growth, development, and an active and healthy life(1). It is often the result of the unavailability of food and/or lack of resources needed to obtain food(1). In 2020, the Food and Agriculture Organization (FAO) reported that globally, 1 in 3 people(2) did not have access to adequate food, highlighting this pressing challenge.

The impact of food insecurity on health varies between high-income countries (HICs) and low-to-middle-income countries (LMICs)(3). In HICs, food insecurity is linked to suboptimal dietary choices, resulting in overweight and obese (OW/OB) weight status and increased vulnerability to diet-related non-communicable diseases (NCDs), such as cardiovascular disease (CVD) and type II diabetes(3–5). In LMICs, food insecurity is more commonly associated with inadequate dietary intake, malnutrition and underweight(3,6). However, many LMICs are undergoing a nutrition transition influenced by economic, demographic, environmental, and cultural dynamics. The full extent of the nutrition transition’s impact on population nutrition remains to be comprehensively evaluated, specifically in the context of food insecurity(3,4,6,7).

As LMICs undergo this nutrition transition, the effects of diet and health factors associated with food insecurity may begin to mirror trends seen in HICs, including increased obesity and NCDs(6). This transition often involves changes in food consumption patterns driven by rapid urbanization, which can exacerbate economic disparities, increase access to ultra-processed foods (UPF), and limit access to nutritious food(8). However, only a limited number of studies have investigated the association between food security and these health outcomes in LMICs. The Philippines serves as a prime example of a LMIC undergoing this nutrition transition(8). Despite economic progress and urban growth, in 2015, 64% of the population was chronically food insecure(9,10). Aging adults in the Philippines are disproportionately impacted, with 52% reporting difficulty accessing food in recent years and significantly more women than males are food insecure(11–13). Specifically, in Cebu province, one of the most economically developed provinces in the Philippines(14), roughly 50% of the population is food insecure(10).

This study examined the relationship between food insecurity and CVD risk among aging women in Metropolitan Cebu, Philippines, an area which includes three of the top eight highly urbanized areas outside the country’s National Capital Region(9). Metro Cebu’s unique position as one of the country’s top three regions for obesity and economic growth(14),(15,16) provides a valuable case for understanding how food insecurity impacts health outcomes in rapidly urbanizing settings(17),(18,19). Specifically, we assessed the association of food security with subsequent CVD risk, represented by body mass index (BMI), waist circumference (WC), and body fat percentage (%BF), as well as clinical outcomes, including incident hypertension and diabetes. Understanding these relationships is crucial for informing public health strategies aimed at mitigating the growing burden of NCDs in LMICs undergoing rapid dietary and lifestyle changes.

## MATERIALS AND METHODS

### Sample

This study used data from the Cebu Longitudinal Health and Nutrition Survey (CLHNS). The CLHNS is a community-based longitudinal study (*n* = 3327) started in 1983 in Metro Cebu, Philippines. Participants came from randomly selected barangays (administrative units), including dense urban, peri-urban, and rural mountain and outlying island settings. The initial purpose of the CLHNS was to explore infant feeding patterns but it was later expanded to include multiple aspects of maternal and child health. Additional details regarding the CLHNS sampling methodology can be found elsewhere(18,20).

We used data from the 2012 (n = 1818) and 2015 (n = 1568) CLHNS follow-up surveys, which were focused on aging in the CLHNS mothers(21). Loss to follow-up occurred primarily due to migration (22). Additional details about sample differences from the previous surveys and the 2012 and 2015 survey have been published(19,21,23). We included participants who were captured in both the 2012 and 2015 survey, yielding an analytic sample of n = 1562. CLNHS data were collected during in-home interviews by trained research staff and in a clinic by nurses or medical technologists. Prior to enrollment, all participants provided informed consent to partake in the study. The research was carried out in compliance with the principles outlined in the Declaration of Helsinki. The study protocol received approval from the Institutional Review Boards at both the University of North Carolina at Chapel Hill (IRB #05-1422 and 11-0064) and the University of San Carlos (no assigned number)(22).

### Food Insecurity

The Radimer-Cornell scale, is a validated assessment tool used previously in the Philippines to represent household and individual food insecurity(24). Our adapted version included 7 items, each measured as an ordinal variable. Participants were asked to reflect on their experiences over the past year, selecting one of three possible responses for each item: 1 (“not true”), 2 (“sometimes true”), or 3 (“often true”) These responses were summed for a maximum score of 21. On this scale, a higher score represents more severe food insecurity, consistent with the interpretation of the literature that the difference between household and individual food insecurity often reflects increased severity of food-related challenges(25–28),(29). We opted to maintain this continuous measure, rather than categorizing participants into discrete categories, as the total score more effectively captured the spectrum of food insecurity severity experienced by participants. The wording of the questions, as predominately “I” statements, were best suited to identify individual food insecurity. The adapted questionnaire and scoring methodology can be found in Supplementary Materials (Table S1).

### Outcome Measures

The following outcome measures, indicative of CVD risk(30–32), were assessed:

### Adiposity Measures

All anthropometrics were collected in triplicate by trained research staff. Using standard techniques, weight was measured to the nearest 0.1 kilogram (kg) using portable Tanita scales, height was measured to the nearest 0.1 centimeters (cm) using portable stadiometers, and WC was measured to the nearest 0.1 cm using standard measuring tapes(33). The mean of the repeated measurements was used as the outcome of interest. Body mass index (BMI) was created using the average weight(kg)/ height(m) squared of each participant. Body fat percentage (%BF) was derived from the Tanita scale’s bioelectric impedance analysis (BIA) feature(34). OB and OW weight status was categorized using World Health Organization (WHO) guidelines as BMI ≥ 30 kg/m^2^ and ≥25 kg/m^2^, respectively. Despite findings suggesting an elevated risk of CVD at lower BMI among Asian population(35,36), we used these thresholds for comparability with other studies(34). High WC was defined as ≥80 cm as suggested by WHO and the International Diabetes Federation (IDF)(37).

### Clinical

Blood pressure was measured in triplicate by trained research staff or medical technologists using appropriate cuff sizes, after a 10-minute seated rest, using OMRON HEM-7211 blood pressure monitors(34). An average blood pressure was created for each participant. We used the WHO definition of hypertension: an average systolic blood pressure ≥140 mm Hg, average diastolic blood pressure ≥90 mm Hg, or taking antihypertensive medications(38). Medication data were reviewed by a trained pharmacist to classify medication types. Type II diabetes mellitus (T2DM) was defined using the mean of 3 measures of glycated hemoglobin (HbA1c) assessed using the NycoCard Reader II(39,40) . We defined diabetes as average HbA1c ≥6.5%(39,40), or taking diabetes medication(39,40). This aligns with the diagnostic criteria by the American Diabetes Association (ADA) and the WHO, and is comparable across studies(22,34).

The incidence of hypertension and diabetes was defined as the development of these diseases between 2012 and 2015 among participants who were disease-free at baseline in 2012. Therefore, the smaller sample size for the logistic regression models is a result of excluding those who had disease in 2012. In Table 1, we represent the prevalence of hypertension and diabetes in 2012 and 2015, while incidence of disease in 2015 is represented in Table 3.

**Table 1.**
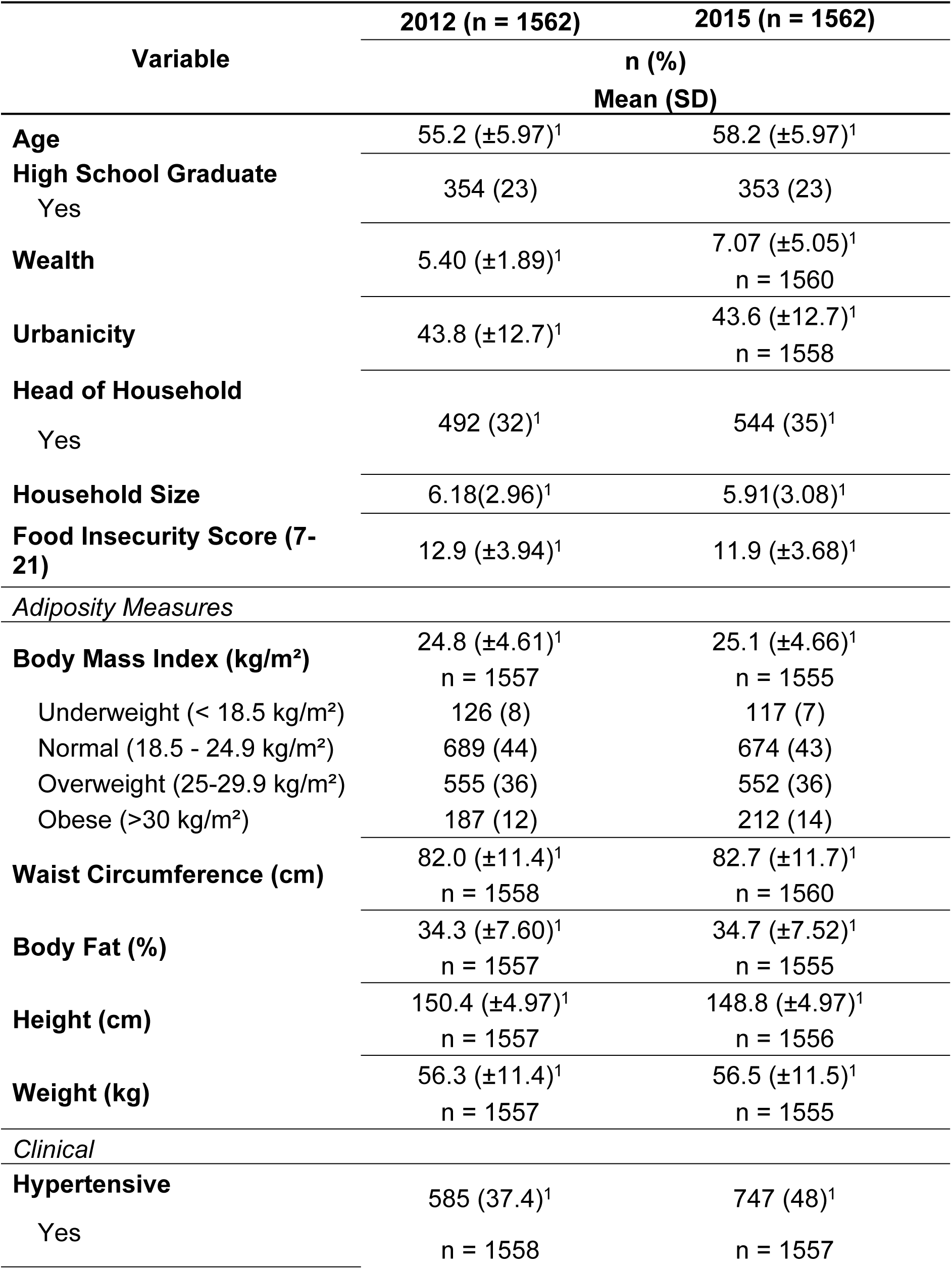
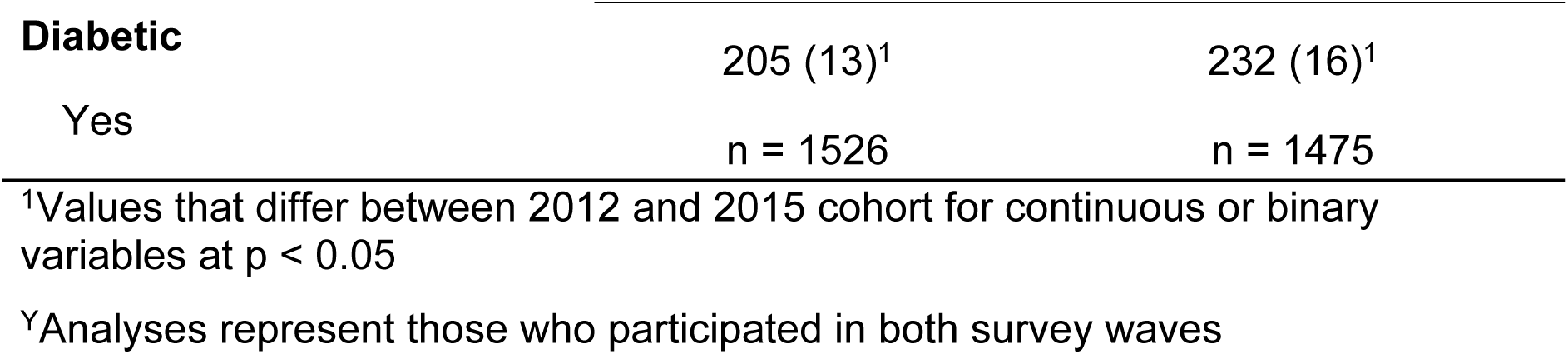
Characteristics of CLHNS women from both the 2012 and 2015 survey (n = 1562)^Y^.

### Covariates

We included age (continuous), women’s status as head of household (HOH) (binary), educational attainment (high-school graduate or not), household size (continuous), household wealth (first principal component of a factor score based on a 30-item household possessions asset index(22),(41)), household size, and urbanicity (continuous, multicomponent index)(18) from 2012. The urbanicity index was based on community surveys with key informants. It aggregates scores from seven components: population size, population density, communications, transportation, educational facilities, health services, and markets(18).

### Statistical Analysis

All statistical analyses were conducted using StataSE 18.5(42).

Differences in participant characteristics across survey years were assessed using paired t-Test for continuous variables and McNemar’s test for binary variables. To assess the association of food insecurity (exposure) with continuous BMI, WC, and %BF, we used multivariate linear regression model as assumptions of linearity were met. For incidence of hypertension, and diabetes, we used logistic regression models. We estimated minimally adjusted models (controlling for participant age) and adjusted models that included age, HOH, household size, and SES variables.

The analysis samples for each outcome varied based on data completeness. For each analysis, we included the participants with data from both the 2012 and 2015 survey (n = 1562) and complete food insecurity data. Missing data were minimal (< 5%) and addressed using a complete-case analysis approach. There were no statistically significant differences in baseline sociodemographic characteristics between women included in the analytic sample (n = 1562) and those lost to follow-up by 2015 (n = 256), as shown in Supplementary Material Table S2 sample sizes are indicated in appropriate models (range n = 975 – 1562).

## RESULTS

### Sample characteristics (Table 1)

Table 1 presents the sociodemographic, dietary, and health characteristics of the CLHNS participants from the 2012 and 2015 surveys. Women ranged in age from 43 to 75 years (mean = 55) in 2012. Household wealth and urbanicity increased, while the average household size decreased from 2012 to 2015. More women reported being HOH in 2015 compared to 2012. Food insecurity scores improved over time, with participants reporting lower food insecurity in 2015. Body fat percentage also increased, in parallel with observed increases in BMI and WC. Consistent with these increases, the prevalence of hypertension and diabetes increased between 2012 and 2015.

### Multivariable models Adiposity Measures

In the age-adjusted model (Table 2), higher food insecurity scores were statistically significantly associated with lower BMI, WC, and %BF. Age was inversely associated with all three anthropometric indices.

**Table 2.**
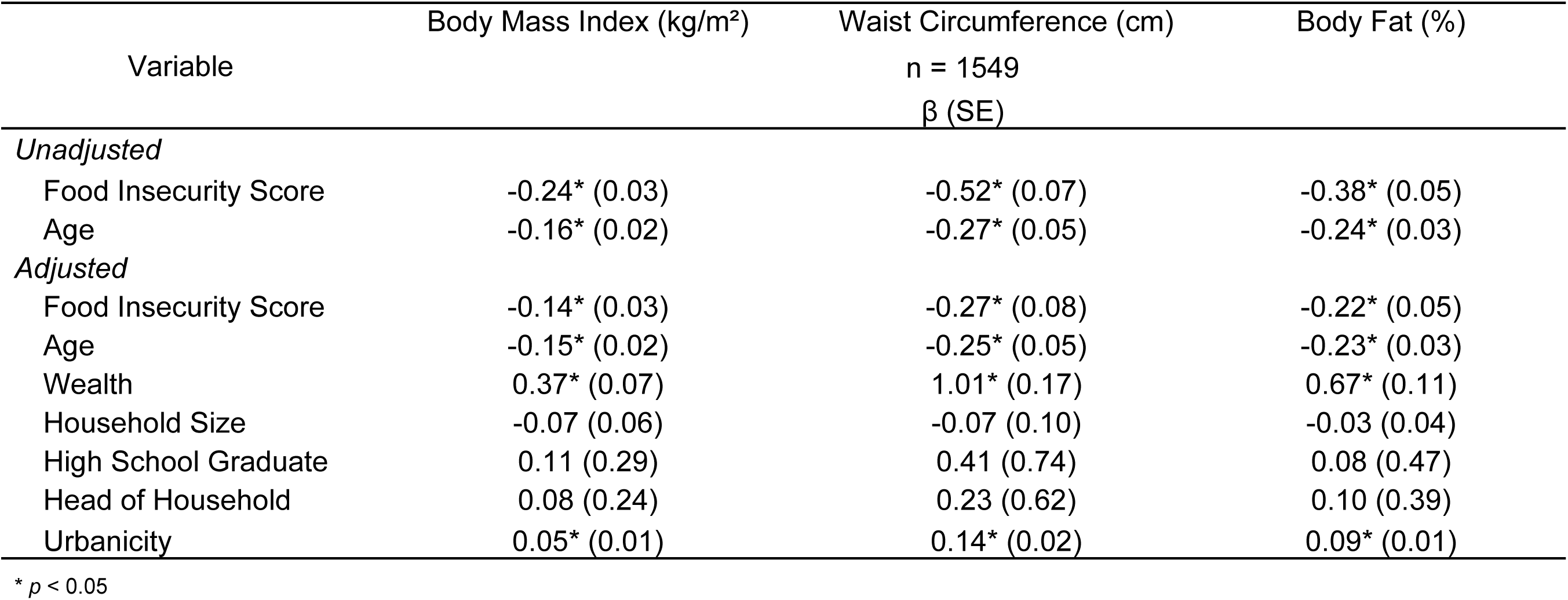
Coefficients and standard error values from unadjusted and adjusted multivariate regression model for adiposity measures by food insecurity score. Unadjusted model is minimally adjusted for age, while the fully adjusted model is controlled for age, wealth, household size, high school education attainment, head of household status, and urbanicity.

| Variable | Body Mass Index (kg/m<sup>2</sup>) | Waist Circumference (cm)<br>n = 1549<br>β (SE) | Body Fat (%) |
| --- | --- | --- | --- |
| *Unadjusted* |  |  |  |
| Food Insecurity Score | -0.24* (0.03) | -0.52* (0.07) | -0.38* (0.05) |
| Age | -0.16* (0.02) | -0.27* (0.05) | -0.24* (0.03) |
| *Adjusted* |  |  |  |
| Food Insecurity Score | -0.14* (0.03) | -0.27* (0.08) | -0.22* (0.05) |
| Age | -0.15* (0.02) | -0.25* (0.05) | -0.23* (0.03) |
| Wealth | 0.37* (0.07) | 1.01* (0.17) | 0.67* (0.11) |
| Household Size | -0.07 (0.06) | -0.07 (0.10) | -0.03 (0.04) |
| High School Graduate | 0.11 (0.29) | 0.41 (0.74) | 0.08 (0.47) |
| Head of Household | 0.08 (0.24) | 0.23 (0.62) | 0.10 (0.39) |
| Urbanicity | 0.05* (0.01) | 0.14* (0.02) | 0.09* (0.01) |

\* $p < 0.05$

After adjustment (Table 2), higher food insecurity scores remained statistically significantly associated with lower BMI, WC, and %BF. Additionally, urbanicity and wealth were positively associated with BMI, WC, and %BF, while age continued to show an inverse relationship with these outcomes.

### Clinical Outcomes

In the age-adjusted model (Table 3), greater food insecurity was significantly associated with a lower diabetes incidence. However, no significant association was observed between food insecurity and the likelihood of developing hypertension in this model. In the fully adjusted model (Table 3), wealth was positively associated with an increased likelihood of developing diabetes. Food insecurity was not significantly associated with the likelihood of hypertension or diabetes in the fully adjusted model.

**Table 3.** Odd ratios (OR) and 95% confidence intervals (CI) for unadjusted logistic regression model for incidence of clinical markers by food insecurity score. Unadjusted model is minimally adjusted for age, while the fully adjusted model is controlled for age, wealth, household size, high school education attainment, head of household status, and urbanicity.

| Variable | Hypertension<br>n = 975 | Diabetes<br>n = 1252 |
| --- | --- | --- |
|  | OR (95% CI) |  |
| *Unadjusted* |  |  |
| Food Insecurity Score | 0.98 (0.95 - 1.02) | 0.93* (0.87 - 1.00) |
| Age | 1.02 (0.99 - 1.04) | 1.01 (0.96 - 1.05) |
| *Adjusted* |  |  |
| Food Insecurity Score | 0.99 (0.96 - 1.04) | 0.99 (0.96 - 1.04) |
| Age | 1.02 (0.99 - 1.04) | 1.01 (0.97 - 1.06) |
| Wealth | 1.07 (0.98 - 1.17) | 1.20* (1.01 - 1.41) |
| Household Size | 0.98 (0.93 - 1.03) | 1.04 (0.95 - 1.13) |
| High School Graduate | 1.08 (0.73 - 1.60) | 0.79 (0.41 - 1.55) |
| Head of Household | 0.88 (0.63 - 1.24) | 0.83 (0.46 - 1.51) |
| Urbanicity | 0.99 (0.98 - 1.01) | 0.01 (0.001 - 0.20) |

\* $p < 0.05$

## DISCUSSION

### Changing demographic and socioeconomic factors over time

The participants in this study exhibited significant changes in the short 3-year period between the 2012 and 2015 survey years. Significant increases in wealth align with broader trends in economic growth and development in Cebu, which may have influenced multiple health outcomes during this period. An increase in wealth may also be indicative of improved purchasing power to afford a consistent and reliable food supply(43),(44). Increased urbanicity may reflect migration to more urbanized communities or changes in the built environment where participants lived. Living in more urbanized communities may affect work opportunities, and increased access to diverse foods, increased motor vehicle use, and healthcare facilities(45). The increased proportion of participants who identified as the HOH from 2012 to 2015 is most likely the result of the loss of a partner. In 2015, 73% of the women who reported being HOH were not living with a spouse and 36% of them were food insecure. Such changes in household dynamics may be indicative of limited economic resources, impacting household purchasing power and food security status(12). Collectively, these factors represent elements of the social and economic environment that influence both food security and our CVD risk outcomes. While treated as confounders in the models, their descriptive trends over time contextualize the broader structural changes that frame the food security landscape in this population. This context is essential for understanding how food security evolves and interacts with health outcomes in transitioning LMICs.

### Adiposity Measures

Studies in HICs have observed a positive association between food insecurity and adiposity, while in LMICs, a negative association has been reported(46), which aligns with the results found in our study. In both the minimally and the fully adjusted models, we observed a statistically significant negative association between food insecurity and adiposity measures. When adjusting for SES indicators, the correlation coefficients between food insecurity and adiposity diminished, but remained statistically significant, indicating that SES partially, but not wholly, explain the relationship between food insecurity and adiposity in our population. In contrast, studies conducted in the Philippines(7) and Vietnam(3), found no significant association between food insecurity and OW/OB status among men and women. These differences may be due to different sample demographics, as aging women may potentially be a more vulnerable group due to limited economic opportunities and differences in our food insecurity assessment. Aging women are also at an increased risk for loss of muscle mass and sarcopenic obesity, necessitating the use of adiposity measures beyond BMI(47,48). Moreover, we used different SES measures, validated in our population and region of interest.

In settings such as the Philippines, food insecurity may correlate with lower anthropometric measures due to limited caloric intake, reliance on basic staples which tend to be low in fat and protein, and constrained access to high-calorie ultra-processed foods (UPF)(45). This divergence may also reflect the unique socioeconomic dynamics within LMICs undergoing a nutrition transition, as wealthier individuals are typically the first to access and incorporate UPF, contributing to an inverse relationship between food insecurity and anthropometric indices of adiposity in poorer populations. The average adiposity measures among our study population should be considered in the context of ethnic-specific adiposity thresholds prior to concluding if undernutrition or overnutrition may be present. While our sample’s average BMI remained below the obesity threshold (≥30 kg/m²), evidence suggests that Asian populations, including Filipinos, may face elevated disease risk at lower BMI thresholds(35,36,49). Research has demonstrated that individuals of East Asian descent tend to have higher body fat percentages at equivalent BMIs than their European counterparts(35,49). Furthermore, our findings reveal that average WC measurements exceeded Asian-specific thresholds, with 56% of participants in 2012 and 59% in 2015 exceeding the IDF cut points for elevated WC(50). Given that WC is strongly associated with metabolic diseases like hypertension and diabetes(49–51), these findings highlight the potential risk of adverse health outcomes in our sample, despite BMI-based classifications that might suggest otherwise. The observed changes in anthropometrics likely reflect a combination of aging, nutrition transition, and socioeconomic factors. However, unmeasured variables such as dietary patterns, physical activity levels, and stress may also influence these outcomes, suggesting the need for further research to disentangle these relationships.

Our study findings also raise critical questions about the differential impact of nutrition transitions across age groups. Our sample represents an older, relatively low-income cohort that may not have been as exposed to dietary shifts brought about by economic growth and the influx of UPF as younger generations(17),(34,52). Limited economic opportunities over the life course, combined with cultural dietary preferences, may mean that this cohort’s exposure to the nutrition transition has been restricted, thereby sustaining traditional eating patterns with lower reliance on processed and Western foods(53). In this context, our findings may reflect both the enduring influence of traditional diets and the delayed or partial impact of the nutrition transition among aging women in the Philippines(17,54). This observation alludes to the importance of examining nutrition transition impacts within specific subgroups, as different populations within LMICs may experience these dietary shifts at different rates and intensities. Therefore, future research should delve into the dynamics of food security, wealth, and anthropometrics in younger cohorts and incorporate dietary data, and potentially other factors along the food security pathway, for a comprehensive understanding of these transitions.

### Clinical Outcomes

The prevalence of hypertension and diabetes significantly increased from 2012 to 2015 (Table 1). This may be partially due to aging and post-menopausal hormonal changes and increasing adiposity from 2012 to 2015, specifically elevated WC and %BF, which is associated with heightened disease risk(55,56). Although our current study covers a short-term period, a previous study using the same cohort found that over a 16-year period, increased adiposity, particularly in the truncal region was associated with increased hypertension(33),

Our study found no statistically significant association of incident hypertension or diabetes with food insecurity scores, before or after adjusting for covariates. Our results were similar to a study conducted in Malaysia, which found no significant difference between food security and hypertension(57). This is relevant to our study due similar geographic location, Asian ethnic background, and influence of Malay cuisine on both countries.

Our findings may be driven by a high prevalence of adiposity, specifically uniformly elevated WC and %BF (Table 1), which may mask the relationship between food insecurity status and disease, as WC is a strong indicator of central adiposity which influences disease risk(49–51). Additionally, this lack of statistically significant findings may, in part, reflect the role of diet not captured by diet-related anthropometric changes. While anthropometric measures such as BMI, WC, and %BF are often direct indicators of disease risk, dietary composition and patterns may also play a direct role in the development of hypertension and diabetes. The traditional Filipino diet, characterized by high-sodium foods such as dried fish (*diang, tuyô, dillis*, *bilad*) with high-sodium condiments (*patis* or *bagoong*), and a heavy reliance on refined carbohydrate staples like white rice, is widespread among individuals with lower SES due to affordability and availability(44,58,59). A diet high in sodium and refined carbohydrates may lead to elevated blood pressure and increased risk of metabolic dysfunction independent of other factors, thereby obscuring a direct link between food insecurity and clinical outcomes in our analysis. Furthermore, coping mechanisms, not captured in the Radimer-Cornell assessment, associated with food insecurity, such as skipping meals, binge eating when food is available, and consuming inexpensive, energy-dense processed foods, can contribute to erratic blood sugar levels and insulin resistance, creating a complex interplay of risk factors that delay or distort the observable relationship between food security and disease incidence(60).

### Challenges and limitations

This study established temporality by using 2012 data to predict incident disease the next 3 years, but yet it does not allow for causal inference in the relationship between food insecurity and CVD outcomes. The adapted Radimer-Cornell food insecurity assessment, although validated in the Philippines(24), may oversimplify the nuanced nature of food insecurity. Our continuous scoring approach better captured food insecurity severity and allowed for easier interpretation but diverges from the traditional scoring method, raising concerns about comparability with studies using the Radimer-Cornell assessment. Given that food insecurity was not the primary focus of the CLHNS, future research should use targeted assessments designed to capture the complex dimensions of food insecurity more effectively. The omission of dietary data as a mediator in this study also limits our understanding of how food insecurity may influence CVD risk. Patterns such as high sodium intake or reliance on carbohydrate staples could independently drive disease risk and should be explored further in future research. Residual confounding is another limitation, particularly due to collinearity between food insecurity and SES variables. Additionally, this study focused on a specific cohort of aging women who were initially enrolled based on childbirth between May 1, 1983, and April 30, 1984, potentially limiting the generalizability of our findings. While attrition due to loss to follow-up, death, or refusal to participate remains a challenge, previous analyses suggest that the 2012 and 2015 participants were not significantly different from the original cohort.

### Strengths

This study designs enables us to establish temporality in describing the association between food insecurity score and CVD risk. Furthermore, it draws upon a well-established data source, focusing exclusively on aging women, which can allow for targeted investigation into the relationship of food security and cardiovascular risk within this demographic. We used wealth and urbanicity indices tailored to our study population, rather than relying on global SES measures. Wealth, which reflects long-term economic stability through assets, provides a more stable SES proxy than fluctuating income, particularly in this aging cohort with limited formal employment.

Similarly, our Cebu-specific urbanicity index better captures regional urban development.(18). By incorporating these tailored SES measures, we were able to better control for these confounders. Furthermore, this study contributes to a scant area of research on the impact of food security and CVD risk in LMICs undergoing a nutrition transition. Aging, post-menopausal women face unique health challenges due to hormonal changes, increased risk of chronic diseases, and socioeconomic vulnerabilities(48). Addressing the intersection of food security and cardiovascular health in this group is crucial for developing targeted interventions and policies.

## CONCLUSION

In this cohort, we found that higher food insecurity scores were associated with lower anthropometric measures, such as BMI, WC, and %BF, reflecting trends observed in LMICs where food insecurity often correlates with undernutrition or limited access to nutrient-dense foods. No significant associations were found between food insecurity and the incidence of hypertension or diabetes. Dietary habits, including high sodium intake and heavy reliance on carbohydrate staples, may independently contribute to disease risk and obscure direct associations between food security and clinical outcomes. Additionally, the relatively short 3-year timeframe of this study may not have been sufficient for disease processes to fully manifest, limiting the ability to detect significant relationships between food security and disease incidence. Future research should address these limitations by employing longitudinal designs over longer periods, integrating detailed dietary data to assess diet as a potential mediator, and refining food security measures that are less collinear with SES indicators. These efforts could help refine policies and interventions for food insecure populations in LMICs undergoing a nutrition transition.

## Data Availability

Most CLHNS data are publicly available on the CLHNS website housed at the Carolina Population Center at the University of North Carolina at Chapel Hill (http://www.cpc.unc.edu/projects/cebu) for researchers who meet the criteria for access to de-identified CLHNS data.

## ACKNOWLEDGMENTS

We would like to express our gratitude to the research staff of the Office of Population Studies Foundation, Inc. of the University of San Carlos in Cebu City involved in the data collection and daily operations to support the CLHNS. The authors are appreciative for the insightful comments by reviewers on the draft of this paper.

